# Source of data for artificial intelligence applications in vascular surgery - a scoping review

**DOI:** 10.1101/2023.10.03.23296506

**Authors:** Katarzyna Powezka, Luke Slater, Michael Wall, Georgios Gkoutos, Maciej Juszczak

## Abstract

**Background:** Applications of Artificial Intelligence (AI) are gaining traction in healthcare, including vascular surgery. While most healthcare data is in the form of narrative text or audio recordings, natural language processing (NLP) offers the ability to extract structured information from unstructured medical records. This can be used to develop more accurate risk prediction models, and to identify patients for trials to improve recruitment. The goals of this scoping review were to determine the source of data used to develop AI-based algorithms with emphasis on natural language processing, establish their application in different areas of vascular surgery and identify a target audience of published journals.

**Methods:** A literature search was carried out using PubMed, EMBASE, and Google Scholar database from January 1996 to March 2023. Following screening, 342 peer-reviewed articles met the eligibility criteria.

**Findings:** Among the 342 articles meeting eligibility criteria, the majority (191) were published after 2020. NLP algorithms were described in 34 papers, while 115 and 193 papers focused on machine learning (ML) and deep learning (DL), respectively. Two-thirds (67.25%) were published in non-clinical journals. The AI-based algorithms found widest application in research related to aorta and its branches (126 articles), followed by carotid disease (85), and peripheral arterial disease (65). Image-based data were utilised in 216 articles, while 153 and 85 papers relied on medical records, and clinical parameters. The AI algorithms were used for predictive modelling (123 papers), medical image segmentation (118), and to aid identification, detection, and diagnosis (103). Only 18 publications aimed to enhance the understanding of vascular disease pathophysiology.

**Interpretation:** Utilisation of different data sources and AI technologies depends on the purpose of the undertaken research. Despite the abundance of available of textual data, the NLP is disproportionally underutilised AI sub-domain.

**Funding:** None.

## Introduction

The concept of artificial intelligence (AI) was first introduced in 1956 (1). According to the Commission Service on digital strategy for Europe, AI systems are software and hardware systems designed by humans that operate in digital and physical realms, gathering and interpreting data from their environment, to make decisions and take actions to achieve complex objectives (2,3). Over the last two decades, an artificial intelligence technology has been increasingly utilised across the healthcare system to improve the quality of patients’ care and efficiency of medical resources and interventions (4). Machine learning (ML) is based on probabilistic and statistical techniques forming complex systems that can iteratively learn from data (5). Deep learning (DL), being a sub-field of ML, is a part of a wider family of artificial neural networks (ANN), defining an architecture that combines several processing layers to learn from data (6,7). There are generally three approaches that ML and DL can utilise: supervised, unsupervised and reinforcement learning (6,8). Natural language processing (NLP) refers to the machine’s ability to identify, process, understand and/or generate information in written and spoken human communications (2). NLP applies computational techniques, that may include ML, or DL, or more basic statistical approaches to analyse speech and text and its tools have been used for information retrieval, classification, text extraction, text summarisation, question answering, and text generation utilising linguistics-focused or statistics-focused approaches, or a combination of these two (9,10). A more extensive explanation of the principles of ML, DL and NLP is included in Appendix 1.

Electronic health records (EHR) allow for constantly growing collection of routine data including clinical notes, radiological reports, radiological and vascular images, physiological and physical metrics, laboratory and histopathology results, and administrative data. There is also an abundance of data derived from large-scale genetic studies and consumer devices such as wearables and smart phones. In many cases traditional statistical approaches fail to parse such large data to provide meaningful results, whereas AI algorithms can effectively handle large datasets and derive information, identifying complex non-linear relationships (11).

Constantly evolving AI technologies are utilised in disease identification, risk prediction and modelling, diagnosis, treatment planning, adverse events monitoring and follow up. Vascular surgery is not different in that respect. However, despite an increasing number of publications, there is a paucity of research exploring the source of information utilised to develop AI algorithms relevant to vascular surgery. To address this, we performed a scoping review that aimed to identify the source of input data utilised to develop AI algorithms with focus on NLP utilisation in vascular surgery. Our secondary goals include assessing application of AI algorithms across vascular surgery and identification of target audience of published results.

## Methods

This scoping review follows the guidelines of the Preferred Reporting Items for Systematic Reviews and Meta Analyses extension for Scoping Reviews (PRISMA-ScR; https://www.equator-network.org/reportingguidelines/prismascr/).

### Data sources

We conducted a literature search using the PubMed, EMBASE, and Google Scholar databases, covering the period from 01/01/1996 to 01/03/2023. No funding source supported this review.

### Eligibility criteria

The predefined eligibility criteria encompassed literature in English or translated to English, published in peer reviewed journals, and met the population, intervention, comparison, and outcome criteria. Reviews, editorials, letters, and case reports were not considered for this review. For the population criteria, the exposure of interest included: aortic disease, carotid disease, peripheral artery disease (PAD), diabetic foot syndrome (DFS), vascular access surgery, arterial trauma, and lower limbs deep vein thrombosis. Articles referring to stroke management, ophthalmology, cardiac surgery, robotic surgery, education and diabetology were excluded. For the intervention criteria, studies were included if they evaluated AI-based algorithms, i.e., ML, DL, and NLP. For comparison and outcome criteria, there were no restrictions.

### Search strategy and selection criteria

We used a combination of medical subjects’ headings, keywords, and other database-specific terminologies selected by a multidisciplinary team composed of AI experts (GG, LS) and vascular surgeons (MJ, MW, KP) to maximise the retrieval of relevant articles, including reference lists of related literature. Artificial intelligence constitutes an umbrella term and the keywords used in the search included “artificial intelligence”, “machine learning”, “deep learning”, “neural network”, and “natural language processing”. Clinical headings and key terms for non-cardiac vascular diseases included “aneurysm”, “carotid”, “peripheral arterial disease”, “revascularisation”, “dissection (artery or aorta)”, “vein”, “vessel trauma”, “venous”, “arterio-venous fistula”, vascular surgical procedures”, “vascular”, “diabetic foot” and “deep venous thrombosis”. We used the Boolean operator “NOT” to exclude irrelevant to the focus of this review conditions, which were “cancer”, “oncological”, “tumour”, “cerebral”, “intracranial”, “heart”, “coronary”, “pulmonary”, “congenital” and “spinal”. The search queries (Boolean expressions) are presented in Figure 1.

**Figure 1.**
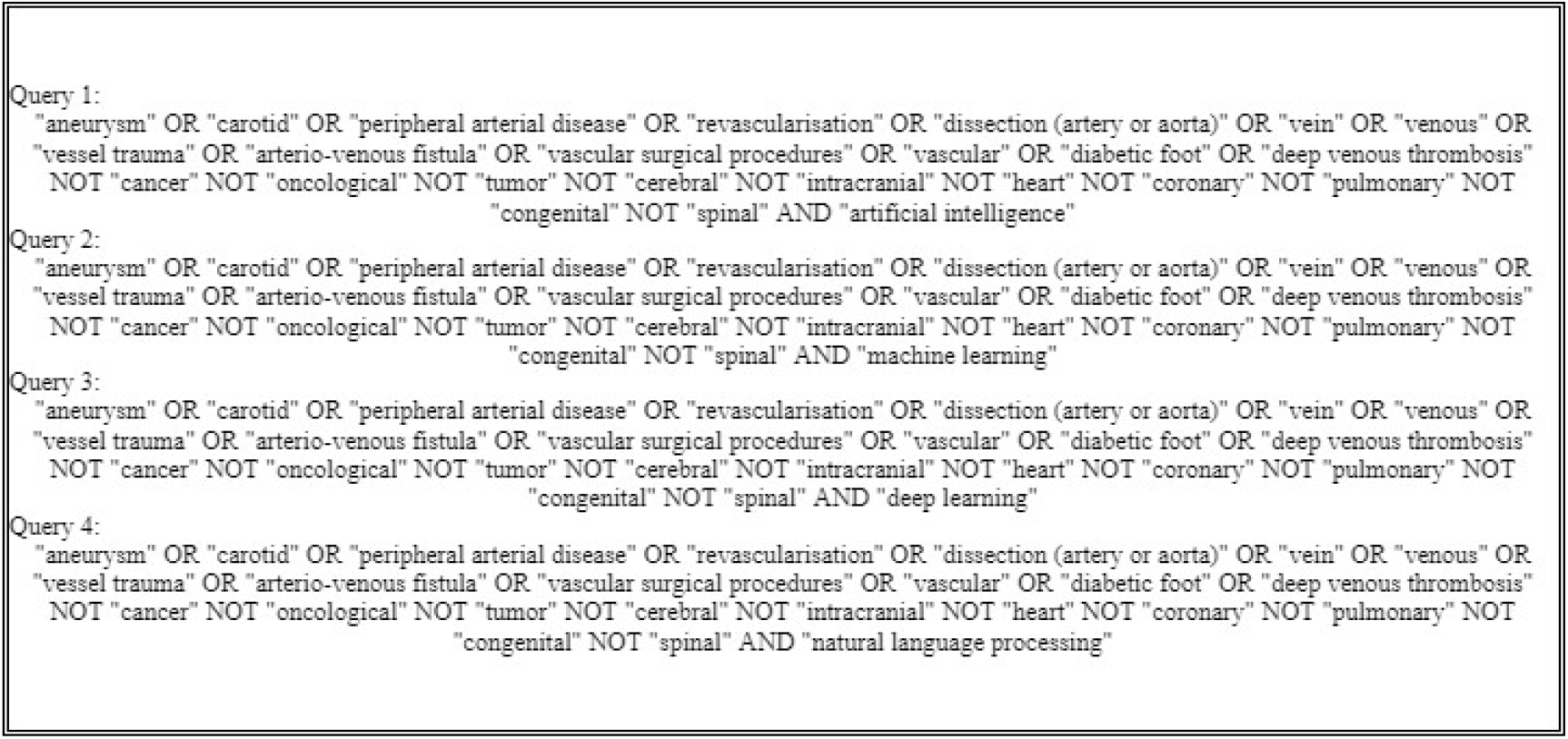
The search queries employed to screen the PUBMED database.

Abstracts were reviewed independently by two authors (KP, LS), and any disagreement was resolved by arbitration (MJ, MW and GG). After identifying titles, full abstracts were reviewed and assessed for eligibility. All citations meeting the eligibility criteria were included in a second round of full-text article screening. Article selection, screening, and data extraction were completed by March 12, 2023. Data were tabulated, summarised, and gaps in AI utilisation in the healthcare system were identified.

### Data analysis

Data that were extracted from the selected articles were stored in an Excel spreadsheet; this included: author, year of publication, journal, title, input data utilised for the AI algorithms development and their application in the vascular surgery.

We divided extracted articles into the following clinical subjects: aortic disease, carotid disease, peripheral arterial disease (PAD), diabetic foot syndrome (DFS), vascular access surgery, arterial trauma, and lower limb deep vein thrombosis. Details of clinical importance of each of these themes are provided in the Appendix 2.

The technologies covered by the umbrella term of AI were classified as machine learning (ML), deep learning (DL) and natural language processing (NLP).

The input data used to develop AI applications were categorised as:

- Image data (derived from computed tomography (CT), magnetic resonance (MR), Xray and ultrasound (US), thermograms, photographs, autofluorescence, microscopic analysis),
- Biological (peripheral blood sample, tissue samples),
- Genome database,
- Medical records (demographic data, medical background, radiology reports, medications, unstructured clinical text, discharge letters, surgical reports, socio-economic status, functional status, type of anaesthesia, medications, surveys results),
- Administrative records (procedure and admissions codes, physician claim codes, hospital volume, surgeon volume),
- Clinical parameters (blood results, heart rate, blood pressure, height and weight, BMI, intraoperative physiological measurements, wound characteristics, ankle-brachial pressure index (ABPI), transcutaneous oxygen pressure, haemodialysis parameters), and
- Devices data (tonometry, doppler sonograms, electrocardiogram (ECG), electromyogram (EMG), ground reaction force, photoplethysmography (PPG), phono-angiography, laser doppler flowmetry, spectrograms).

The intended application of AI-based technologies was classified into the following categories:

- Identification/ Detection/ Diagnosis,
- Prediction,
- Classification,
- Medical image segmentation, and
- Enhancement of understanding of pathophysiological mechanisms underlying vascular diseases.

We summarised the literature in a narrative format for each domain of the vascular surgery focusing on the source of the input data and the intended application of the AI algorithms in the specific areas of the vascular surgery.

### Statistical analysis

Statistical analyses were performed using SPSS (version 21, IBM, Chicago, IL, USA). Data were analysed using descriptive statistics. Results were expressed as counts and proportions/percentage as appropriate.

## Results

We found 584 articles, out of which 342 papers met inclusion the criteria for our scoping review. The article selection is presented in the PRISMA flow diagram (Figure 2). Majority of articles were published after 2020. Articles considering aorta, carotid disease and peripheral arterial disease constituted most of analysed studies (79%).

**Figure 2.**
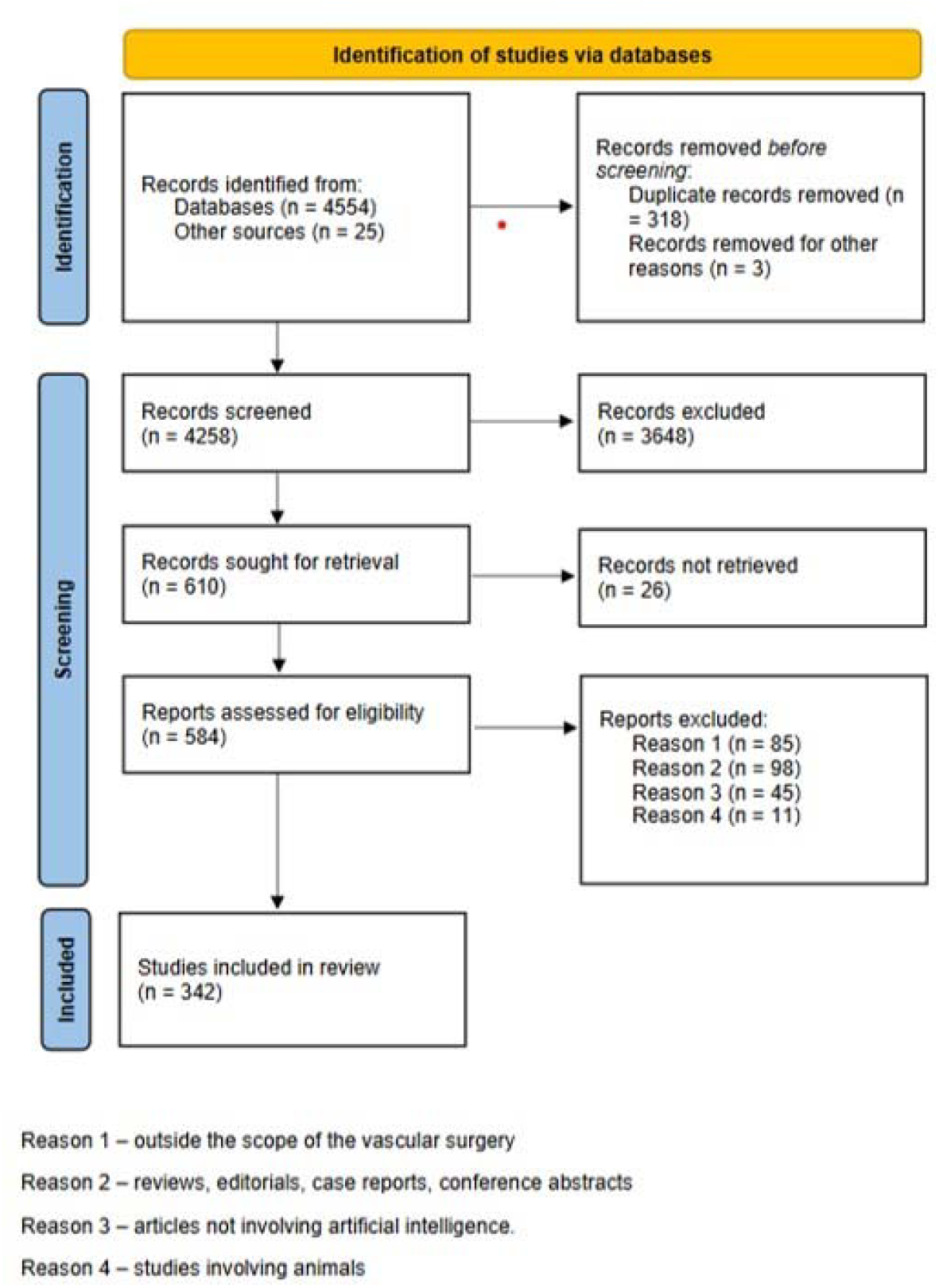
PRISMA diagram from initial literature search to final number of studies included in the analysis.

The most common data sources used for algorithm development were imaging (63·2% of studies), medical records (44·8%), and clinical parameters (24·8%). Biological databases (5·3%), genomic (1·7%) and administrative data (5·3%) were utilised the least (Figure 3). Distribution of AI-based technologies utilised across vascular conditions, based on description of population, intervention, comparison, and outcome criteria of 342 papers included in the review are provided in Table. Application of AI-based algorithms in different vascular conditions is depicted in Figure 4.

**Figure 3.**
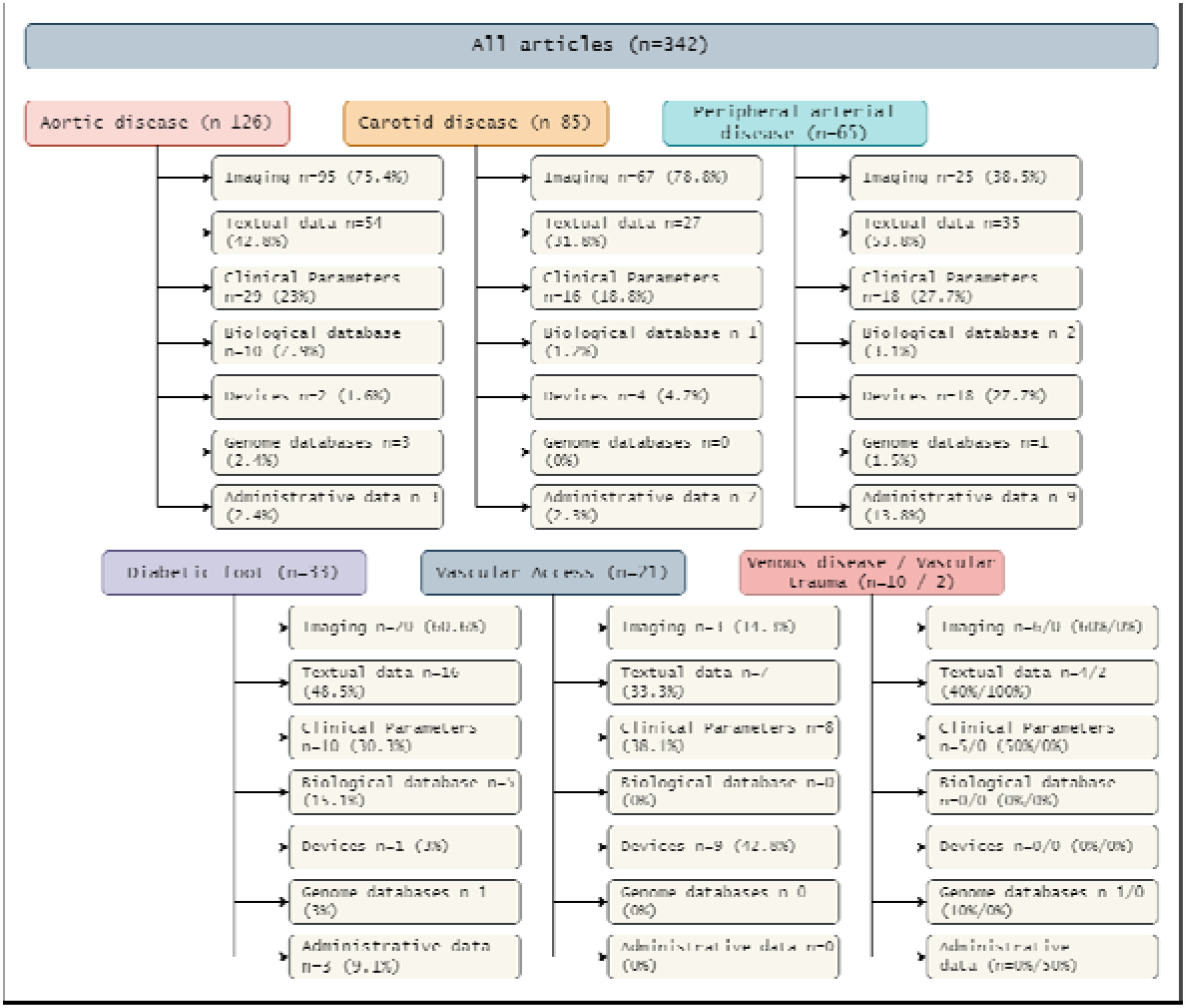
Input data for AI algorithms creation in Vascular Surgery.

**Figure 4.**
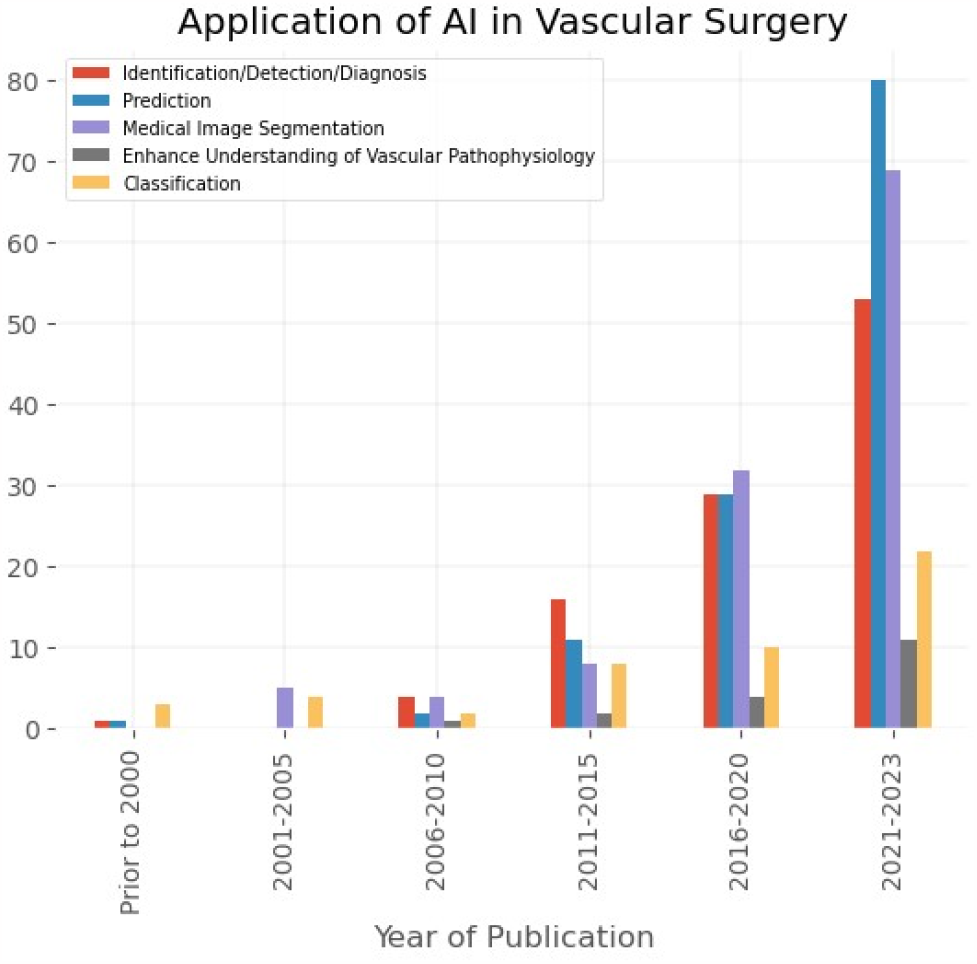
Application of AI-based algorithms in vascular conditions.

**Table.** Criteria based description of included articles.

| Population Data | Articles, n (%) |
| --- | --- |
| Aortic disease | 126 (36.8%) |
| Carotid disease | 85 (24.8%) |
| Peripheral Arterial Disease | 65 (19%) |
| Diabetic Foot Syndrome | 33 (9.6%) |
| Vascular Access Surgery | 21 (6.1%) |
| Venous Disease | 10 (2.9%) |
| Vascular Trauma | 2 (0.6%) |
| **Intervention Data** |  |
| Medical records | 153 (44.8%) |
| Imaging database | 216 (63.2%) |
| Clinical parameters | 85 (24.8%) |
| Biological database | 18 (5.3%) |
| Use of devices | 34 (9.9%) |
| Genome database | 6 (1.7%) |
| Administrative data | 18 (5.3%) |
| **Comparison** |  |
| Machine Learning vs Deep Learning | 42 (12.3%) |
| **Outcome Data** |  |
| Identification/Detection/Diagnosis | 103 (30.1%) |
| Prediction | 123 (35.9%) |
| Medical Image Segmentation | 118 (34.5%) |
| Enhance understanding of pathophysiological processes | 18 (5.3%) |
| Classification | 49 (14.3%) |

| **Population Data** | **Articles, n (%)** |
| --- | --- |
| Aortic disease | 126 (36.8%) |
| Carotid disease | 85 (24.8%) |
| Peripheral Arterial Disease | 65 (19%) |
| Diabetic Foot Syndrome | 33 (9.6%) |
| Vascular Access Surgery | 21 (6.1%) |
| Venous Disease | 10 (2.9%) |
| Vascular Trauma | 2 (0.6%) |
| **Intervention Data** |  |
| Medical records | 153 (44.8%) |
| Imaging database | 216 (63.2%) |
| Clinical parameters | 85 (24.8%) |
| Biological database | 18 (5.3%) |
| Use of devices | 34 (9.9%) |
| Genome database | 6 (1.7%) |
| Administrative data | 18 (5.3%) |
| **Comparison** |  |
| Machine Learning vs Deep Learning | 42 (12.3%) |
| **Outcome Data** |  |
| Identification/Detection/Diagnosis | 103 (30.1%) |
| Prediction | 123 (35.9%) |
| Medical Image Segmentation | 118 (34.5%) |
| Enhance understanding of pathophysiological processes | 18 (5.3%) |
| Classification | 49 (14.3%) |

### Aorta and branches

We found 126 articles (36·8%) focused on aortic pathologies.

NLP technologies were employed in 8 studies (6·4%), utilising unstructured information from medical records (7), administrative and genome databases (1 each) for identification (6), prediction and classification (1 each).

ML algorithms were developed in 42 studies (33·3%) using medical records (19), clinical data (13), biological information (8), genome database (1), imaging data (24) for identification (10), prediction (30), classification (1) and to improve understanding of pathophysiological processes (3) in aortic diseases.

DL technologies were utilised in 76 studies (60·3%) using, as source of information, medical records (23), administrative, and genome database (2 and 1, respectively), clinical parameters (14), biological data (2), imaging and device-derived data (69 and 2, respectively) with the aim of identification (22), classification (4), medical image segmentation (44), prediction (22) and to improve understanding of pathophysiological processes (2).

An external validation of created algorithms was performed in 3 studies. Majority of articles were published in the technical journals (79; 62·7%).

### Carotid disease

We identified 85 articles (24·8%) on extracranial carotid artery disease, majority of which utilised image data to develop AI algorithms (67).

NLP technologies were employed in 5 studies (5·8%), utilising unstructured information from medical records (5) and administrative database (1) for identification (4) and classification (1).

ML algorithms were created in 24 studies (28·2%) using medical records (13), clinical data (10), administrative and biological data (1 in each), imaging- and device-derived data (14 and 3, respectively) and utilised for identification (2), image segmentation (4), prediction (10), classification (7) and to improve understanding of pathophysiological processes (1) in extracranial carotid disease.

DL technologies were utilised in 56 studies (66%) using as source of information medical records (12), clinical parameters (7), imaging and device data (54 and 1, respectively) with aim of identification (5), classification (15), medical image segmentation (48), prediction (6) and to improve understanding of pathophysiological processes (1).

An external validation was performed in 2 papers. Most studies were published in technical journals (71; 83·5%).

### Peripheral Arterial Disease

We selected 65 (19%) articles exploiting AI-based technologies in PAD.

NLP technologies were employed in 15 studies (23·1%), utilising unstructured information from medical records (15), administrative database (6) and clinical data (3) for identification (12) and prediction (4).

ML algorithms were developed in 19 studies (29·2%) using medical records (11), clinical data (8), administrative and genome databases (2 and 1, respectively), imaging- and device-derived data (7 and 6, respectively) and utilised for identification (8), image segmentation (3), prediction (8), classification (2), and to improve understanding of pathophysiological processes (2) in peripheral arterial disease.

DL technologies were utilised in 31 studies (47·7%) using as source of information medical records (9), administrative database (1), clinical parameters (6), biological data (2) imaging- and device-derived data (18 and 10, respectively) with aim of identification (10), classification (11), medical image segmentation (11), prediction (5) and to improve understanding of pathophysiological processes (2).

An external validation of created algorithms was performed in 2 studies. Most studies were published in technical journals (36; 74·2%).

### Diabetic Foot Syndrome (DFS)

Diabetic foot syndrome has come to the focus of AI technologies in the last decade and the volume of published research is growing. We identified 33 studies (9·6%) on diabetic foot syndrome.

NLP technologies were employed in 1 article (3%) utilising unstructured information from genome database for identification and improvement of understanding of pathophysiological processes in the diabetic foot syndrome.

ML algorithms were created in 16 studies (48·5%) using medical records (13), clinical data (8), administrative database and biological data (2 in each), imaging- and device-derived data (6 and 1, respectively) and utilised for identification (2), prediction (9), classification (5) and to improve understanding of pathophysiological processes (1) in DFS.

DL technologies were utilised in 16 studies (48·5%) using as source of information medical records (3), clinical parameters (2), biological data (3), imaging-derived data (13) with aim of identification (6), classification (8), medical image segmentation (4), prediction (2) and to improve understanding of pathophysiological processes (2) in DFS.

An external validation was carried out in 2 studies. Most articles were published in technical journals (23; 69·7%).

### Vascular Access Surgery

Current monitoring and surveillance methods for vascular access remain operator dependent, may be inefficient and may potentially lead to unnecessary interventions. Hence, it is not surprising that amongst 21 (6·1%) identified articles, majority of them (12) were focused on identification and classification of arteriovenous fistula (AVF) stenoses. The input data for ML- or DL-based algorithms were derived from devices used to monitor the function of AVFs, namely photoplethysmography and phono-angiographs. The results from these studies are promising and may lead to constructing point-of-care portable devices for home-based fistula surveillance.

NLP technologies were employed in 4 studies (19%), utilising unstructured information from medical records (4) and clinical data (2) for prediction (3 articles) and improving understanding of pathophysiological processes (2).

ML algorithms were developed in 8 studies (38·1%) using medical records (3), clinical data (5) and device derived data (5) and utilised for identification (5), prediction (3) and classification (1) in vascular access surgery.

DL technologies were utilised in 9 studies (42·9%) using as source of information medical records (2), clinical parameters (1), imaging- and device-derived data (3 and 5, respectively) with the aim of identification (4), classification (3), prediction (2) and to improve understanding of pathophysiological processes (1).

An external validation was carried out only in one study. Most studies were published in technical journals (16; 76·2%).

### Venous Pathology

We have identified 10 studies (2·9%) focusing on utilising ML-based of DL-based algorithms in chronic venous disease (CVD) that is related to daily vascular surgical practice. In 6 of the studies image data derived from photographs, ultrasound, and computed tomography (CT) venograms were employed to develop algorithms. Medical data and clinical information were utilised in 4 and 5 articles, respectively. Only one paper employed genome-based data from Biobank UK to enhance understanding of pathophysiological mechanism underlying CVD. We haven’t identified articles utilising NLP-based technologies. An external validation was performed in 2 studies. Half of the studies were published in technical journals (5; 50%).

### Vascular Trauma

There is a significant paucity of literature on implementation of AI based algorithm in the vascular trauma. We found only two papers, where NLP algorithm or ML-based algorithm were utilised to predict the risks of iatrogenic major vessel injury and failed revascularisation procedures. In both papers the data for algorithms development were derived from medical records and in one also from the administrative database.

## Discussion

We analysed 342 studies on application of AI technologies across different domains of the vascular surgery to assess the types of the data utilised in the development of AI-based algorithms. Most articles were published after 2020 and focused primarily on aortic pathology, carotid disease, and peripheral arterial disease; venous disease, vascular access, and vascular trauma constituted less than 10% of all studies. Despite an abundance of textual data which was used in over 50% of published projects, only one in ten papers described development of NLP algorithms, indicating its underutilisation.

Healthcare leaped forward in quality and complexity during the past two decades, characterised by an exponentially increasing use of information technology, and the vast amounts of data that information technology entails. It was estimated in 2020 that new information generated for every human being per second was around 1.7 megabytes (12). It has been shown that researchers often do not take full advantage of the diversity of data available in the EHR and frequently model development will only use a handful of variables pre-emptively identified by investigators. This limitation in data usage may unduly limit the accuracy of predictive models, especially where complex non-linear interactions are involved (11,13). The textual data in medicine comprises a majority (80%) of information available for analysis (12). However, it is not effective in storing a quantifiable information. A well-designed NLP algorithm would be able to extract a large proportion of quantifiable data from a set of clinical notes without losing it context.

The volume of data and the velocity at which it is being generated exceeds the capabilities of traditional analytical tools. Novel analytical methods are dynamic and utilise advanced statistics, ML, DL, feedback, and NLP tools to mine through the data. These methods provide unparalleled capability in data exploration, analysis, and visualisation; they are robust and capable of seamlessly adapting models over time as new data comes in (12). Rapidly growing medical databanks that include clinical, genomic, imaging, and registries data, will only continue to increase. Thus, the future of medicine is likely to be even more data dependent, with the increasing convergence of medicine and AI technology (14).

The AI potential in healthcare has attracted significant attention from public and private sectors, amongst which Google Health has emerged as a major contributor to the development and deployment of AI-driven healthcare solutions, involving DL and NLP technologies (15). The growing demand for machines to handle complex language tasks, including translation, summarisation, information retrieval, and conversational interactions, has led to development of Large Language Models (LLMs; e.g. ChatGPT, MEDPalm 2) (16). They can be implemented in clinical, educational and research applications. However, since they still are lacking accuracy, they could be only deployed in assisting roles. Their adoption into medicine should be shaped by medical profession with careful clinical governance oversight pending further evaluation and fine tuning (17–19).

Given the context of increasing attention to clinical NLP applications in recent years, we demonstrated that vascular surgery underutilises NLP (18). The narrative data from medical records, clinical parameters, biological and genomic databases were used in 280 (82%) articles, whereas image-based source was utilised in 216 (63%) papers. Despite abundance of narrative data NLP technologies were only utilised in 34 papers (9·9%), whereas ML- and DL-based algorithms were employed in 115 (33·6%), and 193 (56·4%) articles.

There are few reasons that could explain that low utilisation of NLP in vascular surgery. Firstly, vascular surgery is an image-driven specialty; most of decisions including diagnosis, planning of treatment and follow-up are based on various forms of imaging. Secondly, the importance of narrative data is not appreciated in the vascular community. Utilisation of NLP technologies requires labelled data for supervised models, availability of annotators with clinical expertise, and appropriate computational environments. It is technically challenging to meet the hardware and software requirements within existing healthcare IT ecosystems. Additionally, access to patient-level data is controlled by complex information governance regulations, impeding development of efficient NLP algorithms (18). Despite increasing attention to clinical applications of NLP, the proportion of studies using text data in healthcare is low (18,20,21).

NLP holds immense promise in clinical research, with its potential ability to automatically identify cases for clinical studies and trials, drawing from sources like radiology reports. This is particularly important for rare diseases and studies where clinical diagnosis is entered in clinical notes in uncoded manner, and when the precision of diagnostic test is high, but pick up rate is low. Leveraging AI-driven NLP for participant screening using EHR could substantially reduce trials costs and expedite delivery (22,23). This approach has been actively executed by pharmaceutical companies (24,25).

## Conclusions

Literature on AI-based technologies in vascular surgery is dominated by machine learning and deep learning that utilise primarily imaging and structured data. Although NLP holds immense promise, it is underutilised. It is likely that the primary reason for this underutilisation is a deficiency in understanding of this technology among clinicians and lack of adequate expertise. Close collaboration between clinicians and data scientists is mandatory to unlock and utilise NLP in the clinical and research fields.

## Data Availability

All data produced in the present study are available upon reasonable request to the authors
All data produced in the present work are contained in the manuscript

