## Appendix 1 for "Source of data for artificial intelligence applications in vascular surgery - a scoping review"

### Brief description of AI technologies

#### Machine Learning (ML)

In general, the efficiency and the effectiveness of a machine learning solution depends on the characteristics of data and the performance of the learning algorithms. Therefore, selecting a proper learning algorithm that is suitable for the target application in a particular domain might be challenging. The reason is that the purpose of different learning algorithms is different, even the outcome of different learning algorithms in a similar category may vary depending on the data characteristics (1). The availability of the data is considered as a key to construct a machine learning model. One of the key features of machine learning algorithms is their ability to generalize.

Machine learning (ML) approaches can be divided into following four categories (Figure 1):

1. Supervised learning - it uses previously labelled data (training data) to learn its features, so it can perform on similar but unlabelled data. It is used when the certain goals are identified to be accomplished from the certain set of inputs - the task driven process. The most common supervised learning algorithms are classification and regression.
2. Unsupervised learning - it analyses unlabelled data without the need for human interference - the data driven process. This is widely used for extracting generative features, identifying meaningful trends and structures, groupings in results, and exploratory purposes. The most common unsupervised tasks are clustering, density estimation, feature learning, dimensionality reduction and finding association rules.
3. Semi-supervised: It can be defined as hybridisation of the previous two methods, as it operates on the labelled and unlabelled data sets. The ultimate goal of this method is to provide a better outcome for the prediction than that produced using the labelled data alone from the model. This model is used in machine translation, labelling data and text classification.
4. Reinforcement learning - is a type of a machine learning algorithm that enables software agents and machines to automatically evaluate the optimal behaviour in a particular context or environment to improve its efficiency - the environment driven process. It is a powerful tool for training AI models that can help increase automation or optimise the operational efficiency of sophisticated systems (Sarker 2021, Vasilev 2019).

Figure 1. Machine learning techniques.

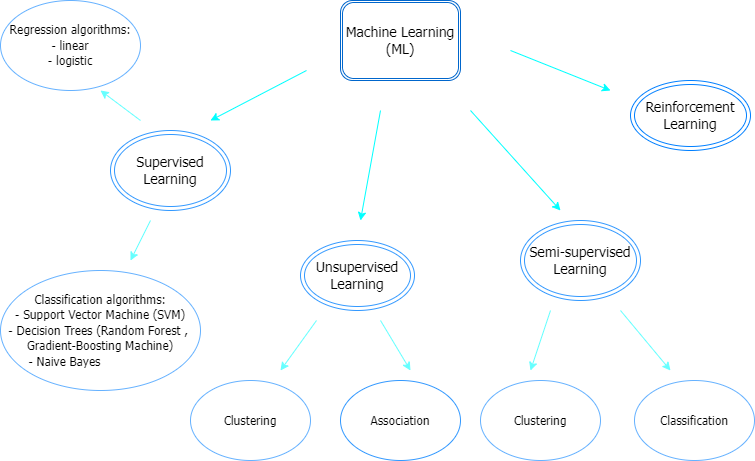

To solve a machine learning problem, the system is required in which a machine learning algorithm is only a part of it. The most important aspects of such a system are a learner, which represents an algorithm, and its choice is determined by the task at hand; training data (labelled and unlabelled); representation of data, which are expressed in terms of chosen features; a goal of the algorithm; and target, which essentially means what was learnt as well as the output of the algorithm (2). A task that is solvable with classic machine learning algorithms requires a thorough understanding and processing training data before deployment. It is important to create, apart from training set, the validation and test sets, as the first is used for tuning the algorithm during the training session and the second for final evaluation and confirmation of the validity of the created algorithm (2).

#### Deep Learning (DL)

Deep learning (DL) is a part of wider family of artificial neural networks (ANN) based machine learning approaches with representation learning. DL provides a computational architecture by combining several processing layers, such as input, hidden and output layers to learn from data. The main advantage of a DL over traditional ML is its better performance in learning from large data sets (Sarker 2021, Yamashita 2018). The most common deep learning networks are:

1. *Mulitlayer Perceptron (MLP)* - it is a class of feedforward artificial neural network (ANN). MLPs models are the most basic deep neural network, which is composed of a series of fully connected layers and each new layer is a set of nonlinear functions of a weighted sum of all fully connected outputs from the prior layer.
2. *Convolutional Neural Network (CNN)* - another class of the DL models for processing data that has a grid pattern, such as images, and designed automatically and adaptively learn spatial hierarchies of features from low- to high-level patterns. It is composed of one or multiple convolution layers that extract the simple features from input executing convolution operations. CNN in general is composed of three layers that are its building blocks (Figure 2):
A. Convolutional layer - being a foundation of CNN, it is composed of a stack of mathematical operations, such as convolution, a specialised type of linear operation. In digital images, pixel values are stored in a two-dimensional (2D) grid, and a small grid of parameters is called a kernel, an optimizable feature extractor. It is applied at each image position, which makes CNN a highly efficient for image processing, since a feature can occur anywhere in the in the image. As one layer feeds its output into the next layer, extracted features can hierarchically and progressively become more complex. The process of optimising parameters such as kernels is called training, which is performed to minimise the difference between the output and the ground truth labels through the optimisation algorithm called backpropagation and gradient decent, among others. Another important part of this layer is the non-linear activation function, which processes the outputs of the linear operations. The rectified linear unit (ReLU) is the most used function.
B. Pooling layer - oversees reducing dimensionality, making it easier to process and requiring less memory. It also helps to reduce number of parameters and makes training faster. Pooling can be divided into maximum or average, depending on the value from the area covered by the kernel - maximum or average, respectively.
C. Fully connected layer (FC) - is one of the most basics layers in a convolutional neural network. It means that each neuron in a fully connected layer is fully connected to every other neuron in the previous layer. FC layers are typically used at the end of a CNN, when the goal is to take features learned by the previous layers and use them to make predictions. The classification procedure gets started at that point.

Along with the above layers, the CNN architecture also encloses activation function, which is the last fully connected layer's activation function is frequently distinct from the others; and dropout layer, which is a mask that nullifies some neurons' contribution to the following layer while leaving all others unchanged and it can be applied to the input vector as well as to the hidden layer. Dropout layer is critical to CNN training because they prevent training data from overfitting (3).

Figure 2. Example of a CNN architecture.

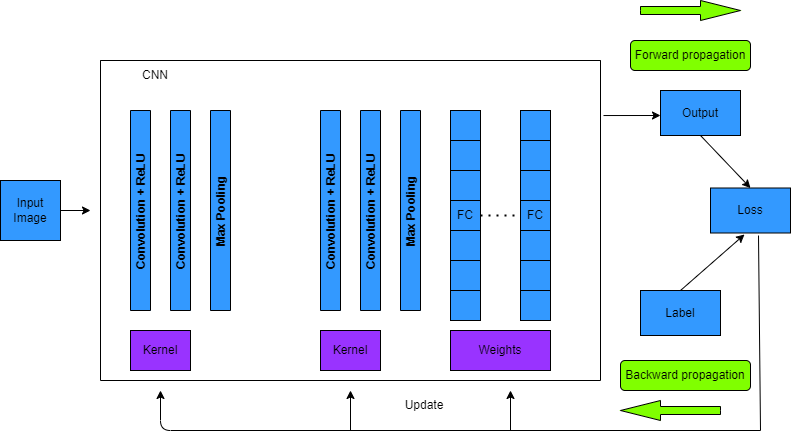

3. *Recurrent Neural Network (RNN)* - is a commonly employed algorithm in the discipline of deep learning, being mainly applied in the field of speech processing and natural language processing (NLP) contexts. Unlike conventional networks, RNN uses sequential data in the network. Since the embedded structure in the sequence of the data delivers valuable information, this feature is fundamental to a range of different applications. Three different techniques of deep RNN were introduced ("Hidden-to-Hidden", "Hidden-to-Output" and "Input-to-Hidden"), that lessens the learning difficulties in deep learning. The biggest disadvantage of RNN is its sensitivity to the exploding gradient and vanishing problems, as during the training process, the reduplications of several large or small derivatives may cause the gradients to exponentially explode or decay. Also, the sensitivity decays over the time, because with the introduction of new inputs, the network stops thinking about the initial ones. This has been addressed with the long short-term memory (LSTM), which offers recurrent connections to memory blocks in the network. Every memory block contains a number of memory cells, which have the ability to store the temporal states of the network. In addition, it contains gated units for controlling the flow of information (2,4). RNN includes less feature compatibility when compared to CNN (4).

Deep Learning approaches in their concepts are similar to those of machine learning, and include: supervised learning, which is employed by RNNS, CNNs, and deep neural networks (DNNs); semi-supervised learning, which is employed by generative adversarial networks (GANs), deep reinforcement learning (DRL), and occasionally by RNNs and LSTMs; and unsupervised learning, which is used by generative networks, dimensionality reduction and clustering, such as auto-encoders, GANs, and RNNs (Alzubaidi 2021).

#### Natural Language Processing (NLP)

Meaning is a fundamental concept in Natural Language Processing (NLP), in the tasks of both Natural Language Understanding (NLU) and Natural Language Generation (NLG) (5). Human language is composed of four major building blocks: phonemes, morphemes and lexemes, syntax and context as presented in Figure 3, and semantics is critical to understanding data and analytics results (6).

Figure 3. Building blocks of language and their applications.

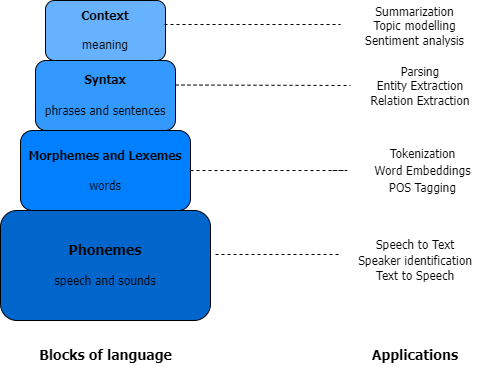

The process of language analysis is being perceived as being decomposable into a few stages, mirroring the theoretical linguistic distinctions drawn between syntax, semantics and pragmatics. The sentences of a text are first analysed in terms of their syntax, which provides an order and structure that is more amenable to an analysis in terms of semantics, that provides a literal meaning. This is then followed by a stage of pragmatic analysis whereby the meaning of the utterance or text is determined (Figure 4) (7).

Figure 4. Stages of analysis in processing a natural language

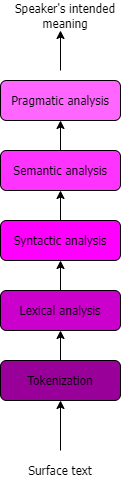

The different approaches used to solve NLP problems fall into three following categories:

1. Heuristics-Based NLP - is a method used in early attempts at designing NLP systems based on building rules for solving a problem at hand. This required that the developers had some expertise in the domain to formulate rules that could be incorporated into a program. An example of designing rules to solve an NLP problem using such resources is lexicon-based sentiment analysis, which uses counts of positive and negative words in the text to deduce the sentiment of the text. Such systems require dictionaries and thesauruses and more elaborate knowledge bases having been built to aid NLP in general and rule-based NLP in particular. The examples of these are regular expressions (regex), that are sets of characters or patterns used to match and find substrings in the text or context-free grammar (CFG) that can be used to capture more hierarchical and complex information that a regex might not (6).
2. Machine learning for NLP - supervised, semi-supervised and unsupervised techniques are applied to textual data. Supervised machine learning techniques, such as classification or regression methods are heavily used for various NLP tasks. One of the classification techniques is Naive Bayes, which relies on Bayes theorem and assumes that each feature in the text is independent of all other features and is commonly used as a starting algorithm for text classification. The goal of any text classification is to learn a decision boundary (linear or non-linear) that act as a separation between different categories of the text. The support vector machine (SVM), which is another popular text classification algorithm can learn both boundaries to separate data points belonging to different classes. Other techniques used in modelling textual data are hidden Markov model (HMM), which assumes that text is generated according to an underlying grammar being hidden underneath a text and each hidden state is dependent on the previous state, and conditional random field (CRF), which performs a classification task on each element in the sequence (6).
3. Deep learning for NLP - neural network algorithms have been increasingly utilised in the language processing tasks. One of the neural networks employed in sequential language processing is recurrent neural network (RNN), which is specially designed to progressively read an input text from one end to another and is equipped with neural units that can remember what they have processed so far. This memory is temporal, and the information is stored and updated with every time step as the RNN reads the next word in the input. The memory problems when processing a long text are mitigated with application the long short-term memory (LSTM) network, which allows to disperse the irrelevant context and focus only on remembering the part of the context required to solve the problem at hand. Convolutional neural network (CNN) has been successfully employed in NLP, especially in the text-classification tasks. The latest entry in the league of deep learning models for NLP are transformers, which are designed to model the textual context but not in a sequential manner and are used for transfer learning, where the knowledge gained while solving one problem is applied to a different, but related problem (4,6).
