## Appendix 2 for "Source of data for artificial intelligence applications in vascular surgery - a scoping review"

### Aortic conditions

Aortic diseases, including abdominal and thoracic aortic aneurysms, represent potentially life-threatening conditions. Guidelines of the European Society for Vascular Surgery have defined management recommendations for the abdominal aortic aneurysms, thoracic aortic aneurysms, and aortic dissections (1–3). Even though large initial maximal diameter is a well-established independent risk factor for AAA rupture, other factors for rupture, including general characteristics of the patients (such as female gender, hypertension, or smoking) and factors related to the aneurysm itself (including AAA growth rate, wall stress, wall stiffness, wall tension, and rapid increase of intraluminal thrombus), have been identified (1,4). Efforts have been made to normalize the aortic diameter to other patient-specific dimensions, including body surface area, height and aortic length, which improved predictions of adverse events compared with using diameter alone (5). In practice, however, the risk of progression and rupture can be difficult to predict, and the decision-making strategy for AAA repair and its management varies widely between countries despite common guidelines from professional societies (4). Expeditious advancements in the machine and deep learning technologies could potentially be utilised in developing a more personalized therapeutic approach considering the patients’ general and clinical characteristics as well as a detailed characterization of aortic pathologies, such as geometry and morphology.

### Carotid disease

The extracranial carotid atherosclerotic disease accounts for up to 15-20% of all ischaemic strokes (6). Cardiovascular disease (myocardial infarction and stroke) accounted for around 31% global mortality around the world in 2017 (7). In a European population of 715 million, 1·4 million strokes occur annually resulting in 1·1 million deaths annually in Europe and is the second commonest cause of death after coronary artery disease (CAD). It is suggested that the number of Europeans living with stroke as a chronic condition may increase by 25% from 3·7 million (2015) to 4·6 million (2035), as a result of the ageing population (8).

### Peripheral Arterial Disease

Peripheral artery occlusive disease (PAD) is a highly prevalent vascular disease associated with high morbidity and mortality risks. In 2010, estimates suggested that >200 million people worldwide were living with PAD. This represented a 23·5% increase since 2000, an increase that is believed to be largely attributable to aging populations and the growing prevalence of risk factors. These figures are thought certainly to almost underestimate the true burden of disease as they are largely based on community-based studies that define PAD on the basis of reduced ankle-brachial pressure index (ABPI) (9). Current diagnosis entails identifying symptoms ranging from classical claudication to rest pain and non-healing wounds. Unfortunately, only 10–30% of PAD patients report stereotypical symptoms, while clinician and patient awareness of PAD is less than 50%. In the absence of unified screening guidelines, PAD remains heavily underdiagnosed (10). Although chronic limb threatening ischaemia (CLTI) is widely believed to be a growing global health care problem, reliable epidemiologic data are extremely limited.

The Rutherford classification system for limb ischemia, which was introduced in 1986 and revised in 1997, is a simple metric that was developed to help guide clinical decision-making. Under these guidelines, more severe classifications (class 4–6) are associated with a 1-year limb loss rate of 20–30% and a 1-year mortality rate of 25%. Amputation, the putative final endpoint for severe limb ischemia, is associated with a 30-day mortality rate of 22% and a 5-year mortality rate of 77% (11,12). Patients with PAD have an increased incidence of major adverse cardiac events (MACE), cardiovascular mortality, and all-cause mortality when compared to patients without PAD. Only 10% of patients with PAD have the classic symptoms of intermittent claudication and approximately 40% do not complain of leg pain, and the remaining 50% have a variety of leg symptoms different from classic claudication. Diminished or absent pulses on physical examination lacks both sensitivity and specificity and may not be sufficient for the diagnosis of PAD. Therefore, identifying patients with PAD is challenging and as such, patients with PAD are often under-diagnosed and under-treated with proven guideline-endorsed therapies (13).

### Diabetic Foot Syndrome (DFS)

Diabetic foot syndrome has come to the focus of AI technologies in the last decade and number of published research is exponentially growing. Diabetes mellitus currently affects 9·3% of the global population (463 million people), 90% of which are afflicted with type II diabetes. This is expected to increase to 10·9% by 2030. Diabetes is the seventh leading cause of death and the single greatest cause of lower extremity amputations. It is estimated that the lifetime incidence of a diabetic foot ulcer (DFU) is between 15% and 25% and that 85% of non-traumatic lower limb amputations are preceded by a foot ulcer. Around 20% of patients with diabetes in hospitals are brought there due to complications from DFU (14). In Europe, 250,000 diabetic patients undergo lower limb amputation with an associated mortality of 30% at one month and 50% at 1 year. Diabetic foot ulceration is associated with markedly increased healthcare costs, decreased quality of life, infection, amputation, and death. The detection of patients at risk of DFU may enable timely intervention to prevent foot ulceration, amputation, and death (15). Moreover, 5-year mortality among patients with a previous limb amputation is more than 50% (16)). Diabetes affects end organs, such as eyes, kidneys, foot, leading to debilitating complications and increased financial burden. Therefore, a lot of emphasis is being put on preventative approach, including prophylaxis, patients’ education in the community and early detection of possible foot complications in the course of diabetes.

### Vascular Access Surgery

Creation of autologous arterio-venous fistula (AVF) represents the gold standard vascular access (VA) for haemodialysis (HD) in patients with end stage renal failure (ESRF) (17,18). Organized efforts to promote autogenous haemodialysis access, most notably the National Kidney Foundation’s Kidney Disease Outcomes Quality Initiative (KDOQI), have resulted in a shift away from prosthetic accesses and tunnelled central venous catheters (CVCs) toward arteriovenous fistulae (AVF). Despite efforts to promote the utilization of autogenous access, nearly half of AVFs created are never used successfully, and 80% of patients initiate haemodialysis with a CVC (19). The latest 2019 Kidney Disease Outcomes Quality Initiative guidelines and 2018 European Society for Vascular Surgery (ESVS) recommend regular physical examination, including auscultation and palpation for vascular access surveillance and monitoring a health practitioner with moderate quality of evidence (17,20). The rationale for surveillance is based on the hypothesis that progressive stenosis can be accurately detected by reduced intra-access flow and increased venous pressure before VA thrombosis occurs (18). The fundamental principle for performing routine vascular access monitoring and surveillance is timely identification and correction of significant stenosis, thus prolonging patency (18,21).

### Venous Pathology

Chronic venous disease (CVD) of the lower limbs is one of the most prevalent medical conditions in the adult population worldwide, representing 1–2% of the healthcare budgets in Western European countries and North America (22). It can lead to chronic venous insufficiency (CVI), with a prevalence of 25% to 40% in females and 10% to 20% in males. Mild to moderate CVI may manifest itself as varicose veins, swelling, lipodermatosclerosis associated with itchiness, burning sensation or pain, hence impacting on patients' quality of life (23). More severe forms of CVI may present itself as venous leg ulcers (VLUs), or in rare cases, have malignant transformation into Marjolin's ulcer (24). Post-thrombotic syndrome (PTS) is a common sequela of deep vein thrombosis (DVT), which is caused by chronic venous insufficiency (CVI), secondary to prior DVT, and can affect up to 50% of patients with proximal DVT within 2 years (25).

### Vascular Trauma

Vascular trauma may occur anywhere in the body and the severity of insult is dependent upon the vessel involved and the structures it supplies. The imaging modality most used within vascular trauma is CT which can be used to identify the location and often the cause of the injury with damage to other structures being visible. Interrogation of this data at present relies on clinical acumen and patient assessment. Vascular trauma often leads to time dependent interventions to gain maximum benefit. Interrogation of imaging or results may be beneficial to the trauma team involved with the patient care. However, there is a significant paucity of literature on implementation of AI-based algorithms in vascular trauma.
